# Overview of COVID-19 Sentinel surveillance in Niger from October 2022 to March 2024

**DOI:** 10.1101/2025.05.22.25328148

**Authors:** Hamidou Lazoumar Ramatoulaye, Adamou Lagare, Oumarou Diallo, Yahayé Hanki, Gado Mahamadou, Moumouni Kamaye, Amina Moussa Boubacar, Abdoulkadiri Aouta Zeinabou, Djamila Diouga, Garba Dangné Mouniratou, Balki Aoula, Elhadji Yacoudima Y. M. Aminou, Sabo Haoua Seini, Julien Poublan, Vincent Richard

## Abstract

**Introduction:** Epidemiological surveillance in community and hospital settings enables active case detection and the implementation of preventive measures to protect the patient’s surroundings.

**Methodology:** The surveillance network consisted of six sentinel sites, five in community settings (Integrated Health Centers - IHC) and one in a hospital setting (National Hospital of Niamey - HNN). Surveillance took place from October 2022 to March 2024. Patients who consented and presented clinical signs consistent with Covid-19 were included. Sociodemographic characteristics, medical history, and clinical signs were collected. A nasopharyngeal swab was taken, and statistical analyses were performed using R software (© R-4.2.1). A 5% significance level was used.

**Results:** The Covid-19 surveillance network included 944 suspected cases, with an overall positivity rate of 1.50%. Males (529, 56%) were more represented than females (415, 44%). The male-to-female ratio was 1.28. Positive cases were more frequent among patients aged 0-5 years, 5-15 years, and over 50 years. Cough (80.04%), dyspnea (46.25%), fever (44.46%), and headaches (38.01%) were the most common symptoms. Correlations were observed between vomiting (p = 0.049), loss of appetite (p = 0.049), cancer (p = 0.0009), muscle pain (p = 0.016), and Covid-19 positivity.

**Conclusion:** The integration of Covid-19 surveillance was effective and facilitated by multisectoral collaboration between community and hospital structures. Maintaining this surveillance network for the SARS-CoV-2 virus will be a major challenge in the future for Niger and, more generally, for African countries.

## Introduction

Coronavirus disease 2019 (COVID-19) by its pandemic scale, cause a significant threat to national public health systems (Ref). For instance, on 30 January 2020, following the recommendations of the Emergency Committee, the World Health Organization (WHO) declared the Covid-19 as a Public Health Emergency of International Concern (PHEIC) and latter on. on 11 March 2020, classified as pandemic (Gupta et al., 2023)). as [1]. As of July 2023, globally 633 million cases of SARS-CoV-2 infections have been reported worldwide, with 6.84 million deaths (Leo et al., 2023).

The rapid transmission and complex structure of the SARS CoV-2 variants has led to the emergence of new variants (Alpha, Beta, Gamma, Delta, and Omicron) that have far exceeded initial expectations [2].

Indeed solid surveillance system is crucial for detecting Covid-19 outbreak and respond promptly for public health actions. Therefore, sentinel system associating syndromic and laboratory surveillance constitute the basic approaches for monitoring respiratory disease outbreaks [3,4],

For instance, many African countries experienced COVID-19 pandemic spread with delay and lower reported cases number, hospitalizations, and deaths. Niger Republic has been reported the first COVID-19 case on 19 March 2020 [5]. The case was firstly reported in the capital city, Niamey, before spreading to other seven administrative regions.. As of December 2023, , 9518 cases and 315 deaths were officially reported (Figure 1, “source owid/covid-19-data”).

**Figure 1.**
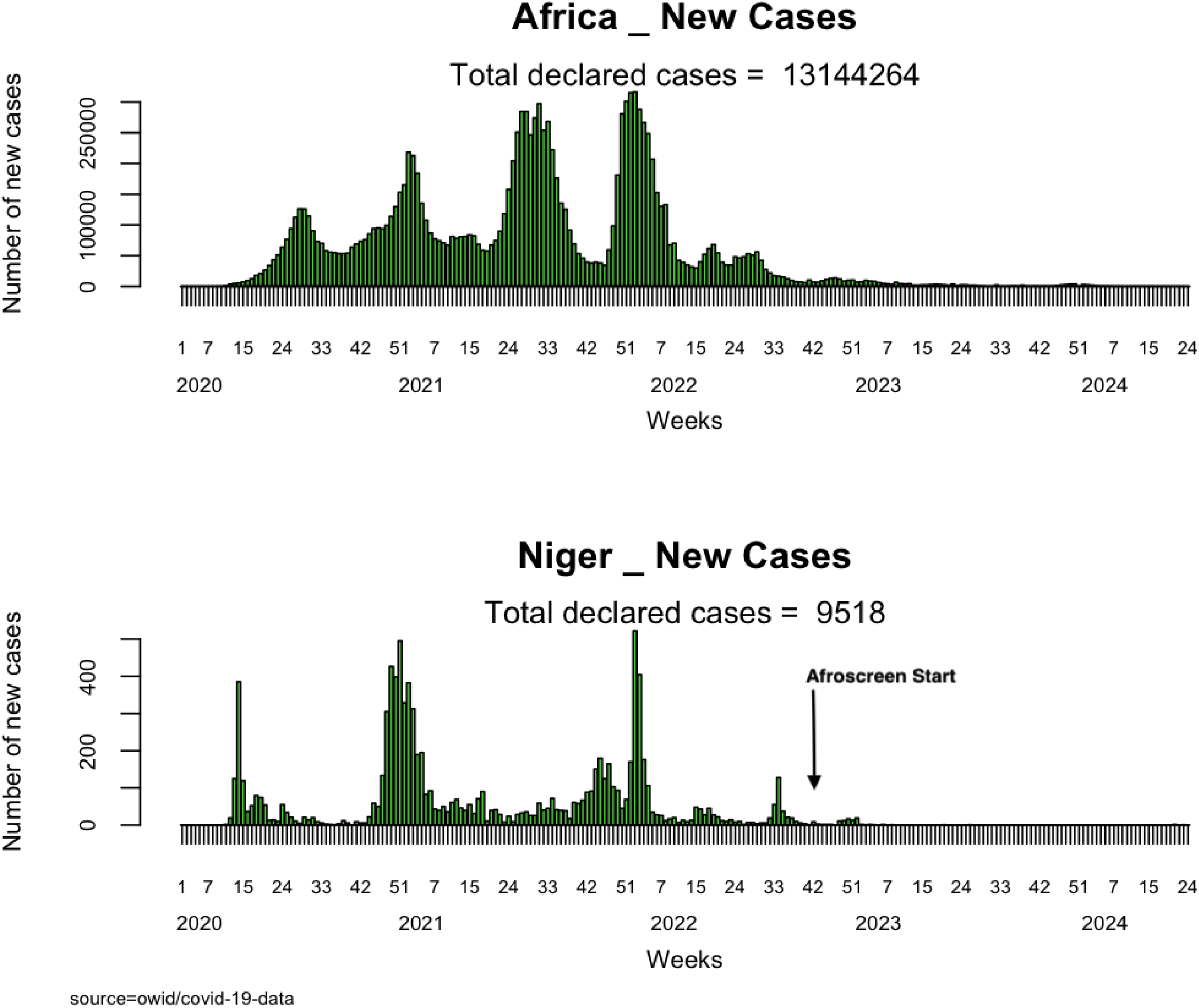
SARS-CoV-2 epidemic curves in Africa and Niger.

This study aims, to understand the main epidemiological characteristics of the Covid-19 cases and monitor the trend of the SARS-CoV-2 circulation. through a sentinel surveillance system.

## Methodology

### Study setting

Niger Republic is a West Africa landlocked continental country, bordered to the north by Algeria and Libya, to the east by Chad, to the south by Nigeria and Benin, to the west by Burkina Faso and to the northwest by Mali. The country covers a surface area of 1,267,000 km^2^and is divided into eight administrative regions including the capital city, Niamey.. characterized by a tropical climate that alternates between four distinct seasons, a long dry season from October to November, a could season from December to February, a hot season from March to May and a rainy season from June to September (Mainassara et. Al, 2015)

### Covid-19 Sentinel surveillance

A Covid-19 sentinel surveillance approaches was set up in six countries of the Pasteur Network including Niger, Tunisia, Madagascar, Central Africa Republic, Cameroon and Ivory Coast through the AFROSCREEN project, a French response initiative, financed by the French Development Agency (AFD) in 2021 in response to these new challenges [6].

In Niger Republic, the system comprises six sentinel sites in Niamey, including one national hospitals namely Hopital National de Niamey (The Infectious Diseases Department and the Pediatrics A and B Departments) and five primary health care centers (CSI Banga Bana, CSI Boukoki 4, CSI Recasement, CSI Lazaret, and CSI Aeroport (figure 2). These health facilities were responsible for enrolling Covid-19 suspected cases based on the established case definition between October 2022 to April 2023.

**Figure 2.**
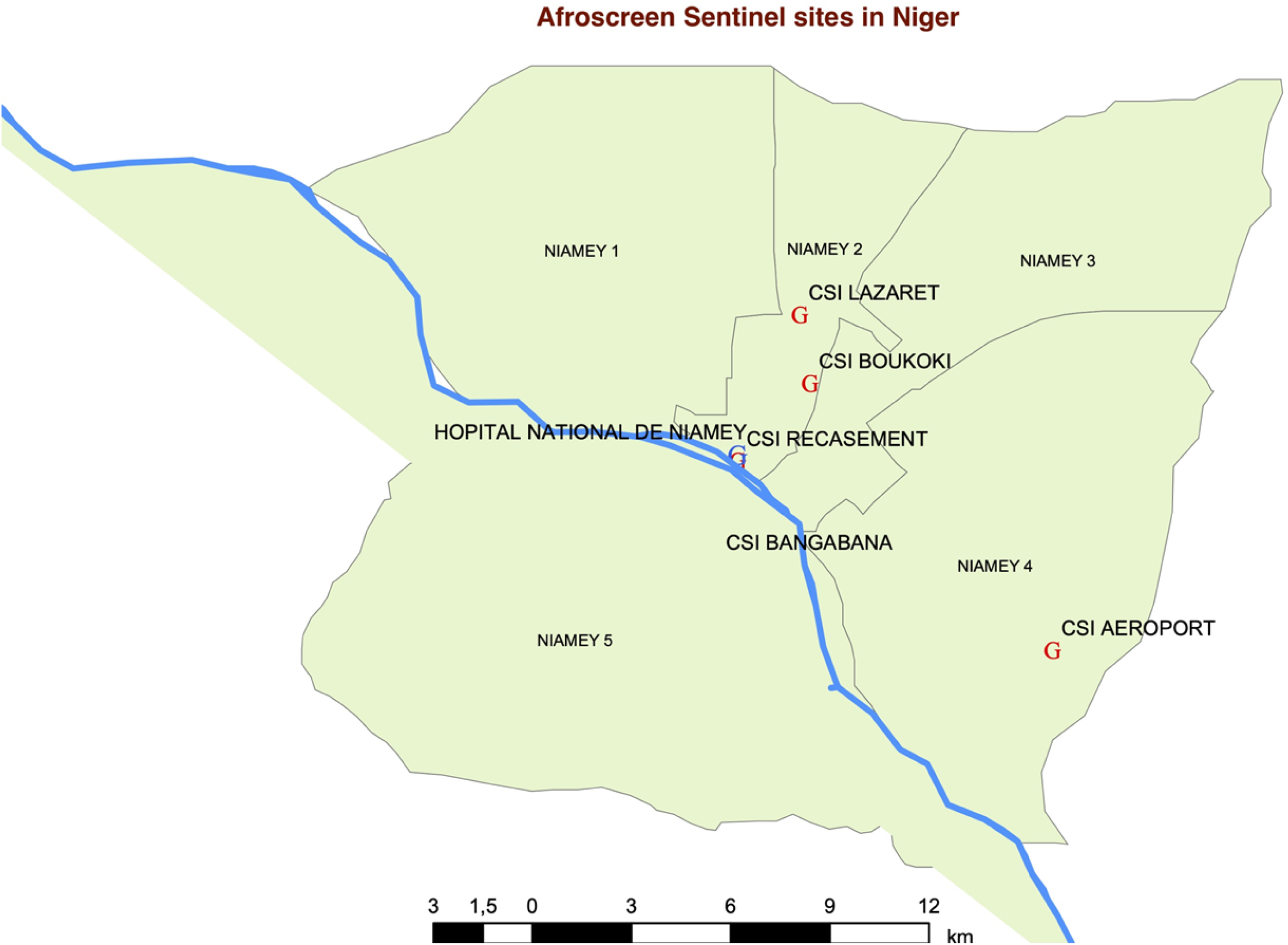
location of sentinel sites in Niamey, Niger.

### Covid-19 case definition

Suspected cases : patient with severe acute respiratory illness: SARI (severe acute respiratory infection) and history of fever, or fever measured ≥ 38°C, and cough, onset within the last 10 days and requiring hospitalization

Confirmed cases : Suspected cases with laboratory confirmation of 2019-nCoV infection

### Laboratory testing

Nasopharyngeal swab samples were collected at each sentinel site from patient’s fulfilling the Covid-19 case definition and transported twice a week at the Centre de Recherche Medicale et Sanitaire, the National Reference Laboratory for influenza and other respiratory viruses. Covid-19 case confirmation were carried out using molecular diagnostic by qRT-PCR technic. Briefly, nucleic acid was first extracted using RADI PREP Swab and Stool DNA/RNA KIT (Ref. MP002) from KH Medical, South Korea. SARS-CoV-2 genome was amplified using RADI Covid-19 detection kit (Ref. RV008) developed by KH Medical and targeting the S and RDRP genes. Positivity was considered for samples with cycle threshold value (ct) ≤ 38.

### Statistical analysis

Data were collected and analyzed with R software (© R-4.2.1). The rRT-PCR result was defined as dependent variables of the study. A Chi-square or Fisher test was performed to compare proportions considering a 5% margin of error.

Logistic regression was performed to determine the clinical signs and risk factors in the form of odds ratios (OR) with their 95% confidence intervals (CI) associated with COVID-19 status.

### Ethical consideration

Ethical clearance was obtained from the National Ethics Committee for Health Research (CNERS) with the reference Resolution N°020/2022/CNERS on 01 juin 2022. Written informed consent was obtained by each participant before collecting blood, respiratory samples and epidemiological data for the intended purpose of this surveillance. Respondents were informed that they had the right to refuse their participation. The information collected was kept confidential. To this end, a code was assigned to each health facility, as well as an identification number for all study participants.

## Results

From October 2022 to May 2024, 944 suspected cases were sampled and among them 14 confirmed cases (Table 1). Global positivity rate was 1.5 %; however, the temporal data showed two periods of Sars-Cov-2 circulation, from February 2023 to May 2023 with a peak on April 2023 (positivity rate = 4.0%) and from October 2023 to January 2024 with a peak in November 2023 (positivity rate = 11.8%). Because of surveillance site modification between the two periods, the comparison of positivity rate could not be interpreted.

**Table 1.** Presentation of the sentinel surveillance activities.

| **Time**<br>**(month/year)** | **Nb of**<br>**sentinel**<br>**sites** | **Sample**<br>**d cases** | **Positive**<br>**COVID-**<br>**19** | **Positiv**<br>**ity**<br>**rate**<br>**(%)** |
| --- | --- | --- | --- | --- |
| 2022 10 | 1 | 12 | 0 | (0.0) |
| 2022 11 | 5 | 70 | 0 | (0.0) |
| 2022 12 | 6 | 139 | 0 | (0.0) |
| 2023 01 | 5 | 29 | 0 | (0.0) |
| 2023 02 | 6 | 53 | 1 | (1.9) |
| 2023 03 | 6 | 185 | 4 | (2.2) |
| 2023 04 | 6 | 50 | 2 | (4.0) |
| 2023 05 | 1 | 90 | 1 | (1.1) |
| 2023 06 | 1 | 48 | 0 | (0.0) |
| 2023 07 | 1 | 60 | 0 | (0.0) |
| 2023 08 | 1 | 26 | 0 | (0.0) |
| 2023 09 | 1 | 30 | 0 | (0.0) |
| 2023 10 | 1 | 24 | 2 | (8.3) |
| 2023 11 | 1 | 17 | 2 | (11.8) |
| 2023 12 | 1 | 22 | 1 | (4.5) |
| 2024 01 | 1 | 20 | 1 | (5.0) |
| 2024 02 | 1 | 28 | 0 | (0.0) |
| 2024 03 | 1 | 41 | 0 | (0.0) |
| **Total** |  | **944** | **14** | **(1.5)** |

In regard of the gender (Table 2), males (529, 56%) were more represented than females (415, 44%).Sex ratio (M/F) was 1.28. The positivity rates were 2% for males (N=10) and 1% for females (N=4) and not statistically different (p-value=0.29). No significant difference related to the period was showed (Table 2).

**Table 2.**
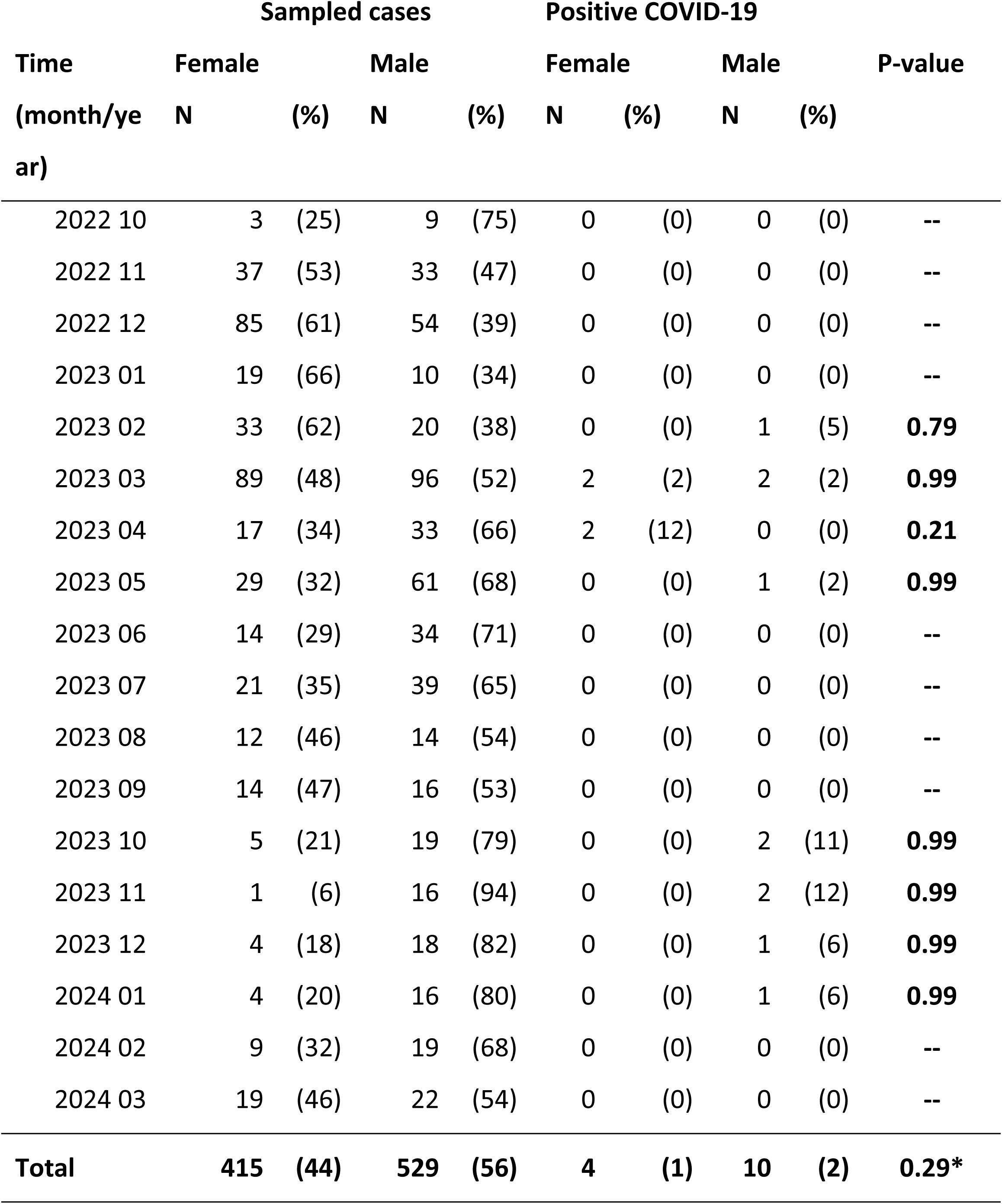
Comparison regarding gender positivity rates.

Considering age group distribution (Table 3), the more represented group was the group of (15-50 years] (54%, n=506), then respectively the group of 50 years and more (21%, n=195), the group of 5 years and less (18%, n=172) and the group of (5-15 years] (8%, n=71). No significant difference was found (p-value = 0.56) for comparison of positivity rate by age groups.

**Table 3.**
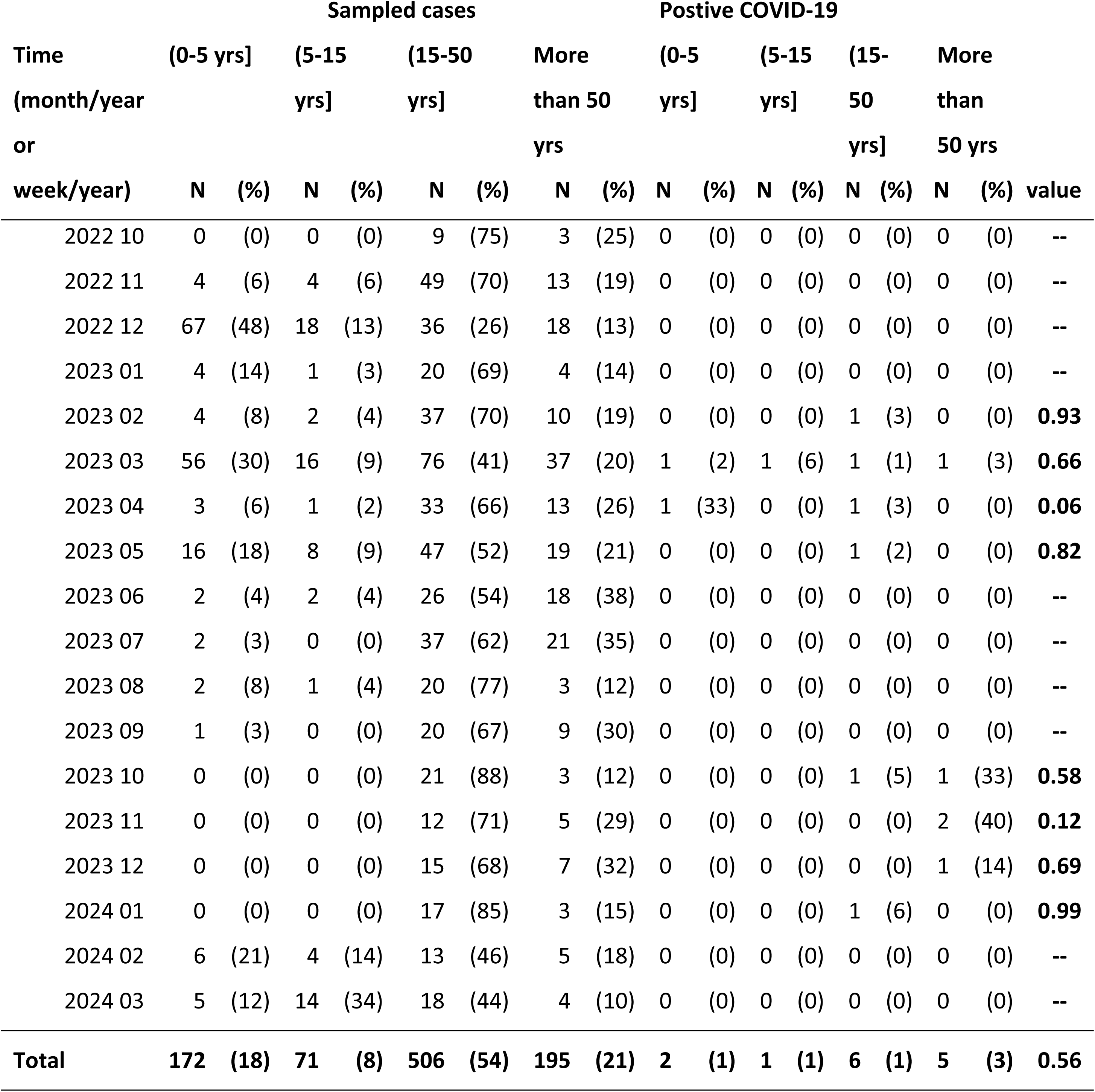
Comparison regarding age group positivity rates.

Considering clinical signs (Table 4), in univariate analysis positivity rates were statistically different for sneezing (p-value <0.01), muscular pains (p-value= 0.016) and vomiting (p-value=0.049). This result was confirmed by the multivariate analysis including clinical variables with p-value ≤ 0.20 (joint pain and cough).

**Table 4.**
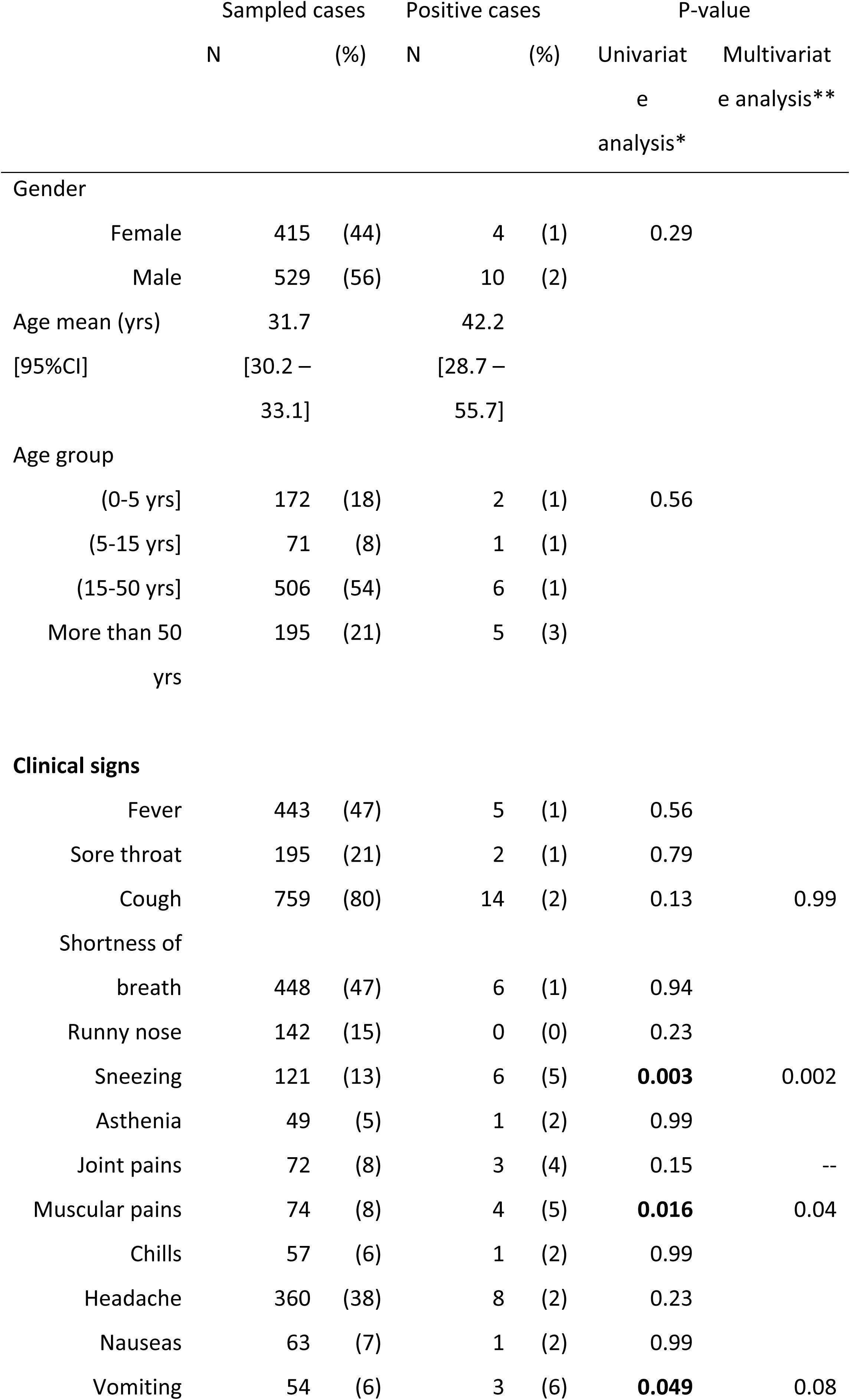
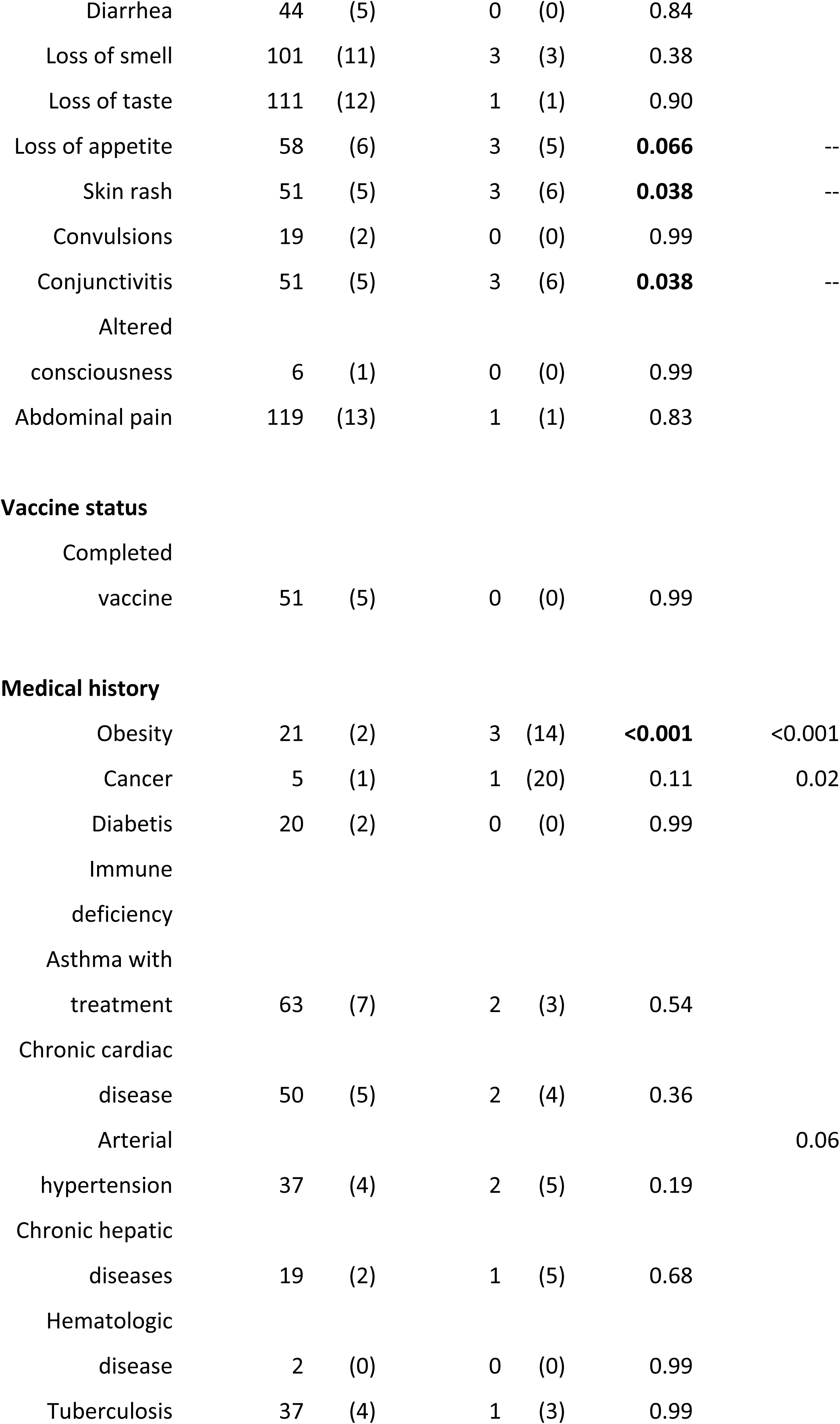
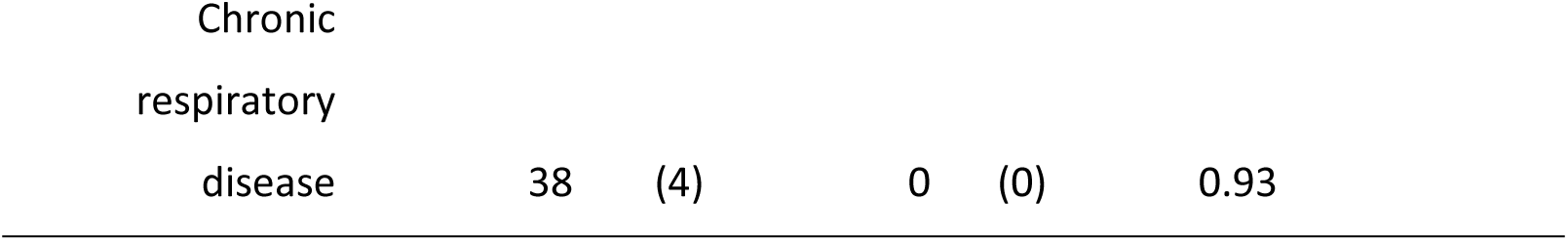
Demographic, clinical features and medical history risks.

Considering medical history and vaccine status (Table 4), in univariate analysis, positivity rate was statistically different for obesity (p-value <0.001). In multivariate analysis, positivity rates were statistically different for obesity (p-value <0.001) and cancer (p-value=0.02) in a regression model including also an history of arterial hypertension.

In regard of the laboratory results all the positive cases tested by sequencing were Omicron variants.

## Discussion

In Niger, sentinel surveillance system has been built over decades as a response to the threat of seasonal, zoonotic (avianH5N1), and pandemic influenza (H1N1 on 2009) and been used for incorporation of COVID-19 surveillance according with the WHO guidelines and recommendations [8].

The COVID-19 has brought significant challenges for genomic surveillance in public health systems in Africa. With the issue of the variants, implementation of sequencing as an additional and sustained tool for surveillance which has been highlighted by Tegally H. et al [9] and Tosta S. el al [10], has been supported by the AFROSCREEN program funded by the French agency for development. Unfortunately, in Niger the sequencing program was implemented on October 2022 after the last wave of COVID-19 (figure 1) when the number of cases was dramatically decreasing.

However, the sentinel surveillance in Niger allowed identification of two periods of SARS-CoV-2 circulation in 2023, from February to May, and in 2024, from October to January, showing the usefulness of sustained sentinel surveillance, if needed. COVID-19 during pandemic waves is what most papers present [11,12], but continuing surveillance is still interesting because it allows us to study SARS-Cov-2 evolution, improve diagnostic assays, and evaluate the effectiveness of current vaccines.

Although not statistically significant, infection rates in people over 50 years and in male were highest between October 2022 and March 2024 in Niger. This is in contrast to what was found in seroprevalence studies in Africa for age classes but conforms with male [13,14]. These results and comparison should be interpreted cautiously due to high heterogeneity between clinical surveillance studies and seroprevalence surveys.

At the beginning of the pandemic, typical symptoms of coronavirus disease 2019 (COVID-19) are fever, dry cough and fatigue and in severer cases dyspnea [15], but in our surveillance data the signs associated to SARS-CoV-2 infection were sneezing and muscle pains. Sneezing was firstly thought to be a much rare symptom, but it has become more common with newer COVID-19 variants and explained [16]. The period of surveillance described here was in relation with the spread of Omicron variants.

Muscle pains have already been describes among the extra pulmonary symptoms in Iran [17] in patients infected with Delta variants, although respiratory presentations of COVID-19 predominate, and it can be considered as an early symptom in COVID-19 patients.

Concerning obesity as a risk factor of infection, it has already been source of interest, mainly for the interaction between COVID-19, obesity and the immune system [18] about severity. Obesity may influence the clinical and immune course of the disease as well as the immune response to vaccines. But our results showed obesity would be a predisposal factor of infection.

Concerning the cancer, our results suggested the higher susceptibility to SARS-CoV-2 infection in patients with cancer, however the number of patients with cancer was a limitation of our results. But the direct effects of cancer on COVID-19 outcomes and hospitalization are already shown [19] and also cancer such as risk factor of infection [13]. The most important limitation of our study came from the limited COVID-19 positive case and the reduction of the number of sentinel site since May 2022. At this time, surveillance was restricted to severe acute respiratory syndromes in hospitals due to decrease of cases sampled in the community health care centers.

## Conclusion

The integration of COVID-19 surveillance was facilitated by existing influenza sentinel surveillance systems, so sustaining permanent global surveillance for respiratory viruses with epidemic and pandemic potential will be a great challenge in future in African countries. Furthermore, the COVID-19 pandemic was the first health major event for which sequencing capabilities have been strengthened in African countries. So, as previously recommended by the Pasteur Network teams in Cameroon [6] sequencing capacities should be sustained and used for detection of new pathogens (pathogen X) among undiagnosed severe respiratory infections and surveillance systems consequently improved in regard of these challenges and these new WGS tools.

## Limits

The implementation of the project’s activities after the Covid-19 pandemic.

## Data Availability

The data are available in a secure database and can be provided if necessary. The contact person is Dr. Hamidou Lazoumar Ramatoulaye, Tel: +227 96134073.

## Funding

« This work was supported by Agence Française de Développement through the AFROSCREEN project (grant agreement CZZ3209), coordinated by ANRS | Maladies infectieuses émergentes in partnership with Institut Pasteur and IRD.

We would additionally like to thank members from the AFROSCREEN Consortium (https://www.afroscreen.org/en/network/) for their work and support on genomic surveillance in Africa. »

## Author contributions

VR, HLR, OD, JP, participated in the conception and design of the study,

HLR, OD, LA, VR participated in the data analysis,

AMB, AKAZ, DD, GDM, BA, YH, GM, MK, collected the field data,

LA, BA have done the laboratory analysis,

HLR, VR, OD, LA, JP contributed to the writing of the text,

VR, JP, SHS, YH GM, MK critically revised the manuscript.

All authors have read and approved the final version of this manuscript.

## Acknowledgements

Our thanks to all the national staffs of the structures implementing the project in the community (CSI Banga Bana, CSI Lazaret, CSI Aéroport, CSI Recasement et CSI Boukoki) and hospital environment (The Infectious Diseases Department and the Pediatrics A and B Departments).

